# Evaluating the AI Potential as a Safety Net for Diagnosis: A Novel Benchmark of Large Language Models in Correcting Diagnostic Errors

**DOI:** 10.64898/2026.02.22.26346832

**Authors:** Ahmed Hassoon, Xiaoyi Peng, Ruxandra Irimia, Anthony Lianjie, Hubert Leo, António Bandeira, Hyun Yi (Jacqualine) Woo, Mark Dredze, Raja-Elie Abdulnour, Kathryn M McDonald, Susan Peterson, David Newman-Toker

## Abstract

**Background:** Diagnostic errors are a leading cause of preventable patient harm, often occurring during early clinical encounters where diagnostic uncertainty is maximal. Large language models (LLMs) have shown potential in medical reasoning, yet their ability to function as a diagnostic safety net, specifically by identifying and correcting human diagnostic errors, remains systematically unquantified. We evaluated whether state-of-the-art LLMs can effectively challenge, rather than merely confirm, an erroneous physician diagnosis.

**Methods:** We evaluated 16 leading LLMs (including GPT-o1, Gemini 2.5 Pro, and Claude 3.7 Sonnet) using 200 standardized clinical vignettes representing 20 high-stakes, frequently misdiagnosed conditions. Models were presented with the full clinical record and an incorrect physician diagnosis. Primary outcomes included the diagnostic correction rate (disagreeing with the error and providing the correct diagnosis) and the ratio of correction to error detection. We further tested model robustness by generating 2,200 variants to assess the influence of demographic (race/ethnicity) and contextual (institutional reputation, training level, insurance) tokens.

**Results:** Diagnostic correction rates varied significantly across models. Gemini 2.5 Pro demonstrated the highest performance, correcting the physician’s error in 55.0% of cases (n=110/200), followed by Claude Sonnet 3.5 (48.5%) and Sonnet 4 (47.0%). In contrast, DeepSeek V3 corrected only 20.0% of cases. Performance was strikingly consistent at the disease level; most models failed to correct errors in syphilis, spinal epidural abscess, and myocardial infarction. Furthermore, several models exhibited confirmation bias (agreeing with the incorrect diagnosis) occurring in 11.0% to 50.0% of cases. Stability across demographic and contextual variants was inconsistent, with some models showing spurious performance shifts based on non-clinical tokens.

**Conclusion:** While top-performing LLMs can intercept approximately half of the human diagnostic errors in high-stakes scenarios, performance is heterogeneous and highly sensitive to non-clinical context. Current models exhibit significant disease-specific gaps and a tendency toward confirmation bias, suggesting that their safe clinical integration requires adversarial, multi-agent workflows designed to prioritize skepticism over baseline agreement.

## Introduction

Diagnostic errors represent a significant challenge to global patient safety. In the United States alone, an estimated 50 to 100 million diagnostic errors occur annually, leading to nearly 1 million cases of serious misdiagnosis-related harm, defined as permanent disability or death.^1^ These errors typically stem from a failure to intervene in patients with dangerous underlying diseases that go unrecognized during initial encounters. Diagnosis is not a static event but a dynamic, iterative process. It begins with clinical suspicion and converges toward clarity through successive testing and medical encounters.^2^ Patients are most vulnerable during the earliest stages of evaluation, particularly in high-volume settings like the Emergency Department (ED).^2–4^ Adverse events among those treated and released after a misdiagnosis can be quantified using the Symptom–Disease Pair Analysis of Diagnostic Error (SPADE) approach, which highlights the precarious nature of early clinical decision-making.^5^

Artificial intelligence (AI), specifically large language models (LLMs), has demonstrated remarkable capability in aggregating and interpreting complex, patient-specific medical data.^6–9^ Leveraging these tools to support diagnosis could fundamentally shift the public health landscape. However, adoption is hindered by concerns regarding training data bias, the lack of prospective validation, ^10^ and the risk of “sycophancy”, where an AI model merely echoes a clinician’s (potentially incorrect) judgment rather than providing a critical second opinion. ^11^

The ability of LLMs to detect and correct human diagnostic errors, particularly during the early “high uncertainty” phase of a disease, remains largely uncharacterized. In this study, we developed a novel benchmark to test LLM in correcting diagnostic errors. We evaluated 16 state-of-the-art LLMs as diagnostic decision-support tools. Our objective was to determine if these models can effectively challenge an erroneous physician decision. A tool that fails to challenge a wrong diagnosis offers limited clinical value and may reinforce cognitive biases like anchoring or premature closure.

## METHODS

### Benchmark Development

We developed a prospective, cross-sectional, clinical vignette-based benchmark to compare the diagnostic performance and error-correction capabilities of LLMs and tested 16 distinct popular models. The benchmark was designed to achieve three primary objectives: first, to quantify the intrinsic ability of LLMs to identify and remediate physician misdiagnoses; second, to compare this corrective performance against the accuracy of the same models in a de novo diagnostic context where no prior physician impression was provided; and third, to determine the extent to which demographic descriptors or other contextual factors (tokens) influence the models’ diagnostic output. The third objective is not part of the public benchmark release. This investigation focused on a high-fidelity clinical scenario reflecting early diagnostic decision-making, simulating an initial visit where a physician has formulated an incorrect diagnosis. This framework allowed us to assess whether AI can function as a true diagnostic safety net, identifying discrepancies between its computational impression and a clinician’s preliminary assessment.

### Case Development and Standardization

Building on our prior work on estimating the type, rate, and clinical presentations associated with diagnostic errors, and informed by our institutional diagnostic safety dashboard, malpractice claims data, and voluntary diagnostic error reporting, we identified 20 most misdiagnosed conditions that are associated with subsequent patients’ harm.^3–5,12–21^

For each of the 20 diseases, we developed 10 diagnostic error cases to generate a total case vignette bank of 200. The intent was for these vignettes to be based on real-world diagnostic error scenarios; however, to ensure equal disease coverage, some cases had to be constructed based on prior diagnostic error experiences and knowledge of experts. The 200 curated vignettes derived from previously published medical literature (n=110), our team’s prior misdiagnosis research (clinical or malpractice cases) (n=50), or were constructed by clinical experts (n=40) (Figure S1).

All cases met the following inclusion criteria (as determined by clinical experts): early visits with clear temporal progression of symptoms and findings, available laboratory and imaging results (if applicable), enough information to consider the correct underlying diagnosis, documented wrong diagnosis, and true confirmed diagnosis. To ensure clinical integrity and realism of the physician-generated cases (n=40), two physicians (S.P. & R.I.) conducted reviews of each case to ensure clinical validity. The distribution of cases by generation method was well distributed across different diseases. Finally, each vignette was transformed into a standardized clinical chart template, encompassing the chief complaint, history of present illness, past medical history, medications, allergies, social and family history, physical examination findings, and initial clinical course. At the end we produced a database of standardized charts of 200 patients who have been misdiagnosed. The final database represents the *origin* cases for case variants (see next section). Supplementary Appendix Figure S1 depicts the flow of case generation. Supplemental attachment S1 provides a sample case.

### Case Variants

To evaluate the robustness and reliability of the models, we generated 2,200 variants of the original 200 cases. The primary variant set consisted of de novo cases where the incorrect physician diagnosis was removed, leaving only the clinical findings. Subsequent variants introduced specific “token” modifications to assess the impact of non-clinical factors. We created four demographic versions for each case, varying only the patient’s race or ethnicity (White, Black, Hispanic, and Asian) while maintaining identical clinical data. Further variants modified the healthcare setting (highly reputable academic center versus an unknown community hospital), the training level of the physician (board-certified specialist versus resident physician), and the patient’s insurance status (comprehensive versus basic). This systematic perturbation of the input data allowed for a granular analysis of how social and institutional descriptors might introduce or mitigate bias in AI-driven diagnostic reasoning. In total, excluding the *origin* cases, we created 2,200 new variants cases of the *origin* cases (Fig. S1).

### Model Selection and Testing Protocol

We evaluated 16 widely used LLMs: DeepSeek (V3, chat; R1, reasoner), Anthropic (Claude Sonnet 3.5 - 20241022, Sonnet 3.7 - 20250219, Sonnet 4 - 20250514, Claude Opus 4.1 - 20250805), Google (Gemini 2 Flash, Gemini 2.5 Pro), Open AI (GPT-4o-2024-08-06, GPT-4.5-preview-2025-02-27, GPT-5-2025-08-07, GPT-o1-2024-12-17), X-AI (Grok-2-1212, Grok-3-beta, Grok-4-0709), and Amazon (Nova Pro v1.0). All models were accessed through their respective API endpoints using their most recent available versions as of August 2025.

Because we curated data of misdiagnosed cases, we tested the models against all cases with the assumption that an average physician misdiagnosed every single case; therefore, average physician performance was set to zero. We used zero shot prompting to mimic a decision support tool workflow for diagnostic safety net establishing model baseline performance, and studies showed that zero shot prompting is comparable for chain of thoughts prompting for clinical diagnostic tasks.^22^ For the primary task, models were prompted with: “Do you agree with the physician diagnosis? Yes, or No. If No, suggest the most likely diagnosis.” In the de novo condition, models were simply asked for the most likely diagnosis. To ensure data integrity, each case-model interaction was conducted in a new session to prevent context contamination. We standardized API parameters, setting the temperature to 0.0 or the minimum possible value, and allowed for unlimited reasoning tokens where applicable.

### Response Classification

For the *origin* cases, we classified each LLM response into three categories: (1) agreed with the incorrect diagnosis, (2) disagreed with the incorrect diagnosis and suggested the correct alternative diagnosis, and (3) disagreed with the incorrect diagnosis but suggested wrong alternative diagnosis. For the de novo diagnosis, each LLM response was categorized into suggested correct diagnosis or wrong diagnosis.

### Statistical Analysis

Primary analyses were conducted using Python (version 3.12.9). For each model, we categorized responses into three types: agreement with the incorrect diagnosis, disagreement with the correct alternative provided, and disagreement but with an incorrect alternative. We calculated the diagnostic error correction rate and the ratio of correction to error detection. Reliability was assessed through the coefficient of variation (CV) and the signal-to-noise ratio (SNR) across token variants. We developed a composite reliability score, normalized on a scale of 0 to 1, where 1 represented maximal performance stability across all contextual perturbations. Computational efficiency was also recorded as the average processing time per 200-case batch.

### Benchmark Availability and Usage

The benchmark and sample dataset, including code script and visualization, are publicly accessible on GitHub (see data sharing statement). Researchers can also expand the number of cases and/or diseases by uploading their own diagnostic error cases to run the benchmark and compare it to other models.

## Results

### Overall Diagnostic Performance and Correction Rates

Our benchmark was design to evaluate the diagnostic accuracy of 16 LLMs and their ability to correct the initial physician’s incorrect diagnosis. The overall diagnostic disagreement and correction accuracy of the LLMs varied significantly. When provided the physician’s incorrect diagnosis (Table 1 and Fig. 1.A), DeepSeek V2 had the highest diagnostic disagreement but the lowest diagnostic correction rate and the lowest ratio of correction to error detection at 0.23. Gemini 2.5 Pro had the highest number of corrected diagnoses, (55.0% of the cases) with ratio of correction to error detection at 0.83, closely followed by Sonnet 3.5 (48.5%) and Sonnet 4 (47.0%) for corrected diagnoses and 0.78, 0.86 ratio of correction to error detection, respectively. In this same condition, several models demonstrated lower performance, with Nova Pro (24.0%) and DeepSeek V3 (20.0%) recording the lowest rates of corrected diagnoses.

**Table 1.** Models’ Errors detection and errors corrections.

| # | Model | Diagnostic Disagreement<br>n(%) | Diagnostic correction<br>n(%) | Ratio of correction<br>to error detection |
| --- | --- | --- | --- | --- |
| 1 | DeepSeek V3 | 176 (88) | 40 (20) | 0.23 |
| 2 | Nova Pro | 148 (74) | 48 (24) | 0.32 |
| 3 | GTP-4o | 82 (41) | 49 (25) | 0.60 |
| 4 | Grok 2 | 111 (56) | 69 (35) | 0.62 |
| 5 | GPT-4.5 | 131 (66) | 73 (37) | 0.56 |
| 6 | GPT-o1 | 138 (69) | 74 (37) | 0.54 |
| 7 | Gemini 2 Flash | 126 (63) | 75 (38) | 0.60 |
| 8 | GTP-5 (HR) | 134 (67) | 76 (38) | 0.57 |
| 9 | Grok 3 | 112 (56) | 76 (38) | 0.68 |
| 10 | DeepSeek R1 | 120 (60) | 82 (41) | 0.68 |
| 11 | Sonnet 3.7 | 130 (65) | 91 (46) | 0.70 |
| 12 | Grok 4 | 142 (71) | 94 (47) | 0.66 |
| 13 | Opus 4.1 | 136 (68) | 94 (47) | 0.69 |
| 14 | Sonnet 4 | 109 (55) | 94 (47) | 0.86 |
| 15 | Sonnet 3.5 | 125 (63) | 97 (49) | 0.78 |
| 16 | Gemini 2.5 Pro | 132 (66) | 110 (55) | 0.83 |

**Figure 1.**
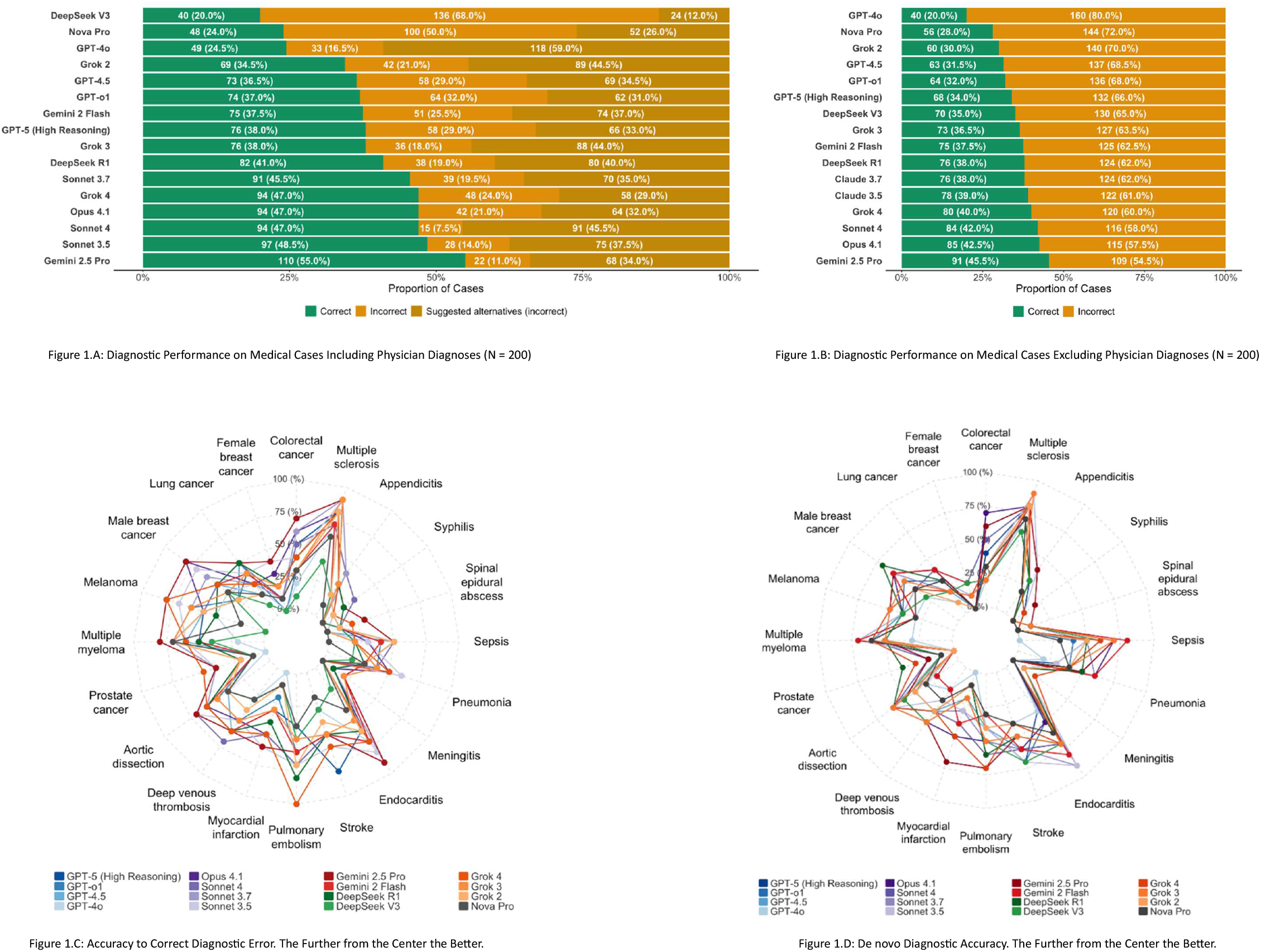
LLMs Performance to Correct Diagnostic Errors

Interestingly, some models correctly identified an error in the initial diagnosis but suggested an incorrect alternative. This may indicate some models’ diagnostic disagreement is not due to better knowledge. This was very evident as models that corrected the diagnoses had high ratio of correction to error detection, while models with lowest performance had the lowest ratio of correction to error detection (Table 1).

In a de novo diagnosis, when LLMs were not provided with the physician’s incorrect diagnosis, overall accuracy was lower across all models. In this condition, Gemini 2.5 Pro is still the top performer followed by Opus 4.1. The models with the lowest performance in this scenario were Nova Pro (28.0%) and GPT-4o (20.0%) (Fig. 1.B).

### Disease-Specific Performance Patterns

We further analyzed the diagnostic accuracy for specific disease categories using radar charts to visualize models’ performance. When an initial incorrect diagnosis was included (Fig. 1.C), performance was inconsistent across different conditions. Most LLMs demonstrated high accuracy for appendicitis, colorectal cancer, and multiple sclerosis. Conversely, accuracy was generally lower across the models for conditions such as syphilis, spinal epidural abscess, myocardial infarction, and prostate cancer. A similar pattern of variable performance was observed when no initial diagnosis was provided (Figure 1.D). While there is variability among models’ performances by specific diseases, poor performance is common across all models for specific diseases. Interestingly, models from smaller to larger share similar diagnostic correction profiles, just at a different percentage (Fig. 2 and Table S1).

**Figure 2.**
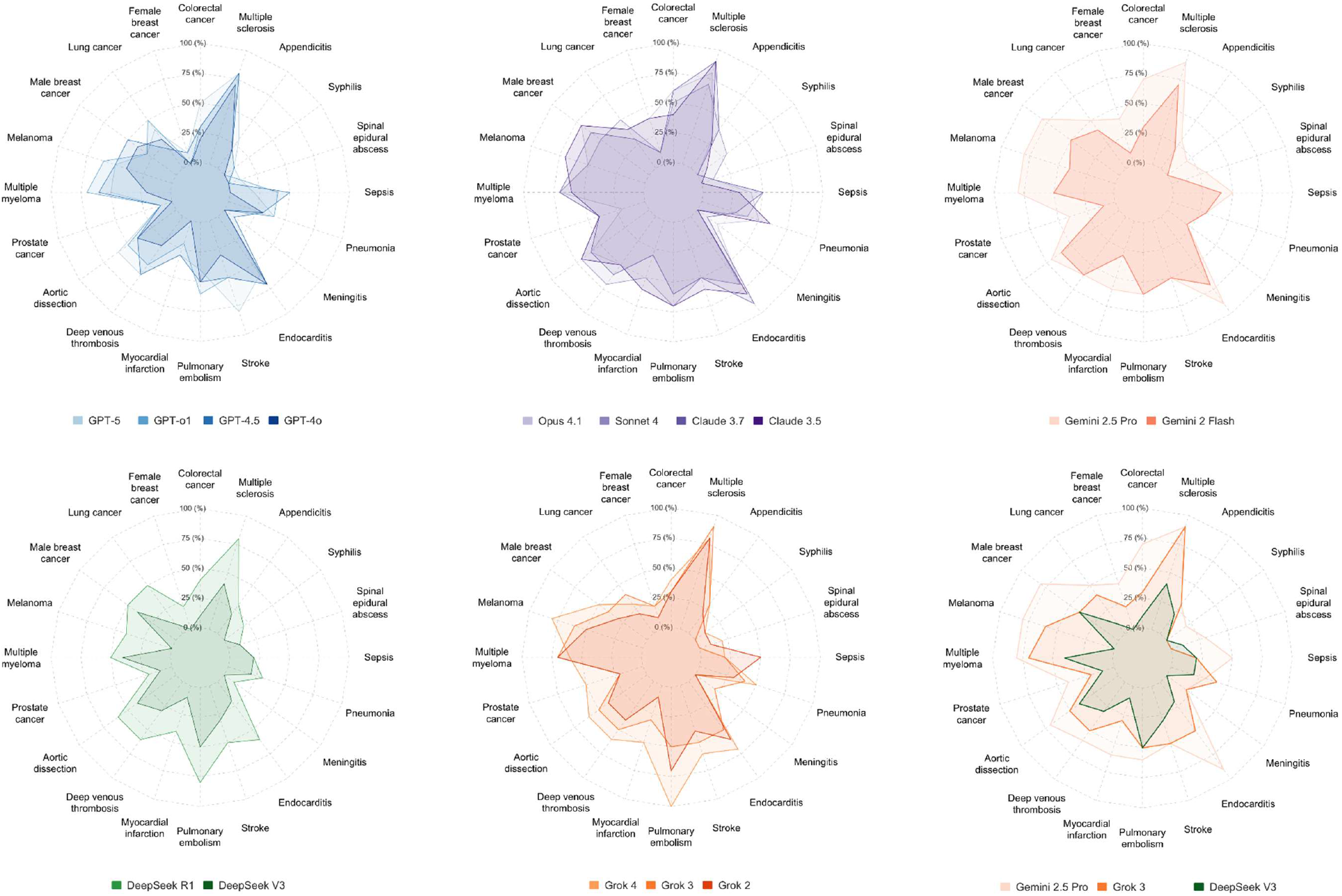
LLMs Performance to Correct Diagnostic Errors by Creator and Versions

### Influence of Tokens Variations

Insertion of demographic, institutional, training, and insurance-related tokens altered LLM performance in correcting diagnostic errors. Across models, average performance shifts were observed, but the direction and magnitude varied by token. Certain tokens, such as “Community-Trained” and “Black,” tended to improve error correction rates slightly above baseline for multiple models, while others, including “Basic Insurance” and “Community Hospital,” were associated with small performance declines (Fig. 3 and Table S2).

**Figure 3.**
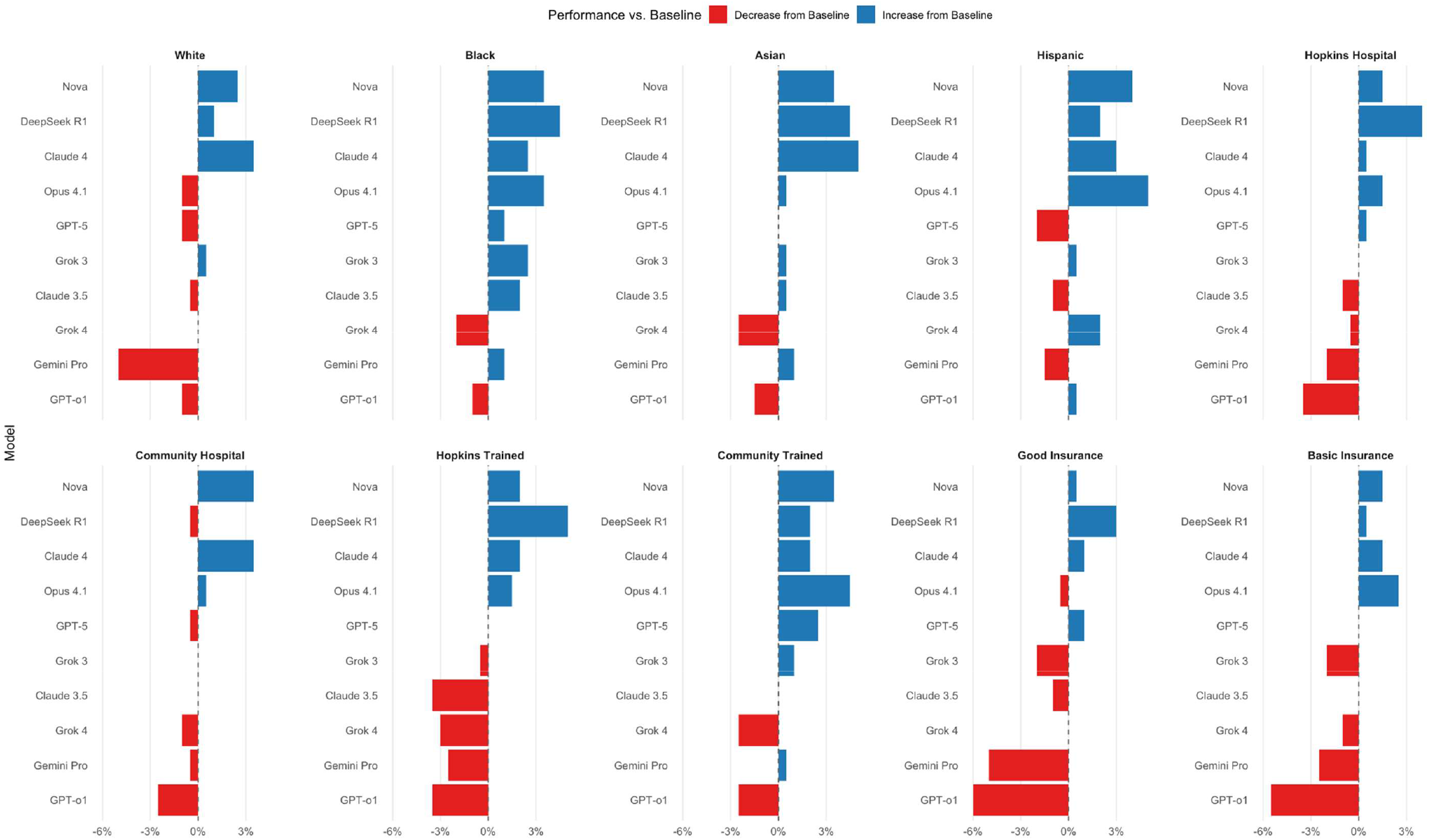
LLMs Performance Stability to Correct Diagnostic Errors by Token Variants Introduction

The extent of performance variability differed substantially across models. Claude Sonnet 4 demonstrated the highest stability, with limited fluctuations across tokens and the highest composite reliability score (Table S3). In contrast, GPT o1 exhibited the greatest instability, with wide swings between best- and worst-performing token contexts. These findings indicate that, in general, models are highly sensitive to token insertions, raising concerns about their robustness in diverse clinical settings.

### Computational Efficiency and Prompting Stability

In a test of performance stability by different prompting strategies (short versus comprehensive), we tested the highest (Gemini 2.5 Pro), mid (Grok 3), and lower (GPT-4o) performing major models. Gemini and Grok were statable under different prompting approaches. GPT-4o’s performance increased slightly using comprehensive prompting approach; however, it was not enough to change the overall rank of the model. Therefore, we recommend using the shorter simpler prompting strategy. As for models’ computation time, the average runtime for every 200 cases significantly varied among models. Longest time was 248 minutes for GPT5, while Grok 3 scored the lowest time at 2.1 minutes. It is important to consider the runtime variability when selecting a model based on performance versus runtime, or when orchestrating different models as they vary significantly (Table S2).

## Discussion

In this benchmark, the evaluation of 16 large language models (LLMs) applied to diagnostically challenging cases where a physician had already committed an error, the best-performing model corrected the misdiagnosis in 55% of instances. Given the high uncertainty setting of early clinical presentations, where clinicians are most susceptible to premature closure, this degree of error interception is potentially clinically significant. These findings suggest that when deployed as a “second reader” with an explicit mandate to challenge initial impressions, LLMs may prevent a substantial portion of downstream harm otherwise solidified by an incorrect diagnostic label. However, performance was heterogeneous across models and disease states, exhibiting sensitivity to non-clinical token changes, such as demographics, institutional reputation, and insurance status, which underscores that clinical utility will depend on reliability as much as peak accuracy.

Across models, the radar plots show strikingly similar shapes: high correction for entities such as appendicitis, colorectal cancer, and multiple sclerosis, and low correction for syphilis, spinal epidural abscess, myocardial infarction, and prostate cancer. This convergence suggests that model family or number of parameters is not the dominant driver of performance at the disease level. Two explanations are plausible and not mutually exclusive: task-intrinsic difficulty at presentation, or knowledge and representation gaps. Some “hard” diagnoses (a) are underrepresented in public web corpora relative to their clinical salience or (b) require up-to-date procedural knowledge and decision rules typically encoded in guidelines or EHR context rather than in general text. However, recent medical-agent benchmarks emphasize uneven coverage across task types (query versus action) and highlight how formatting, tool use, and strict output constraints expose model brittleness even when “knowledge” is present.^23^ Complementary surveys of medical LLM agents likewise point to hallucination management, multimodal integration, and cross-department orchestration as open problems that can depress performance on complex cases.^24^ Together, these observations favor a disease-wise difficulty landscape that today’s models all “see” similarly; larger or newer models shift the curve upward but rarely change its shape. The visual alignment in our radar plots, across unrelated model families, supports this interpretation. The convergence of profiles argues for disease-targeted audits and mitigations rather than generic scaling alone. Contemporary guidance on LLM agents and clinical explainability emphasizes systematic agentic evaluation (plan-critique-verify), retrieval to shore up knowledge coverage, and transparent rationales tailored to high-risk use (Phase 3–4 decision support), all of which can be layered atop the same base models without changing their weights.^25–27^ Sequential diagnosis systems (e.g., MAI-DxO) provide one implementation path, improving both accuracy and cost efficiency when the agent is allowed to seek missing information, precisely the information early visits often lack.^28,29^

Our analysis revealed that models were more effective at error correction when confronted with a specific, but wrong, physician diagnosis than at de novo diagnosis on the same chart. Presenting a concrete hypothesis appears to act as a foil that triggers adversarial reasoning (“Do I agree?”) and narrows the search space. Without that foil, the model must amortize attention across a much larger hypothesis set and, absent iterative querying, can disperse probability mass thinly. This aligns with our finding that de novo diagnosis underperforms the “agree/disagree → suggest an alternative” framing is consistent with emerging agentic evaluations. Systems that explicitly seek disconfirming evidence and structure a challenger role (e.g., MAI-DxO’s “Dr. Challenger”) both improve accuracy and curb anchoring on early hypotheses, precisely the cognitive step our prompt elicits when it asks the model to take a stance on the clinician’s diagnosis.^29^ Similar conclusions are drawn in broader commentaries on LLM agents in healthcare, which argue that multiagent orchestration (plan, critique, verify) is pivotal for diagnostic reliability rather than static, one-shot answering.^27^ Practically, it suggests that clinical deployments should prefer skepticism-first workflows over free-form “what is the diagnosis?” prompts.

While average performance shifts associated with demographic and contextual tokens were modest, several models exhibited meaningful instability. Sensitivity to race, insurance type, or site prestige, while clinical content remained constant, signals fragility in the inference pathway and raises critical fairness concerns. Our composite reliability metric highlights this disparity: while some models remained stable across tokens, others fluctuated significantly, a vulnerability that may be amplified in complex agentic systems involving multi-agent handoffs. Our token sensitivity findings argue for predeployment bias audits that explicitly test demographic and context perturbations, with thresholds for acceptable variance and escalation policies when outputs cross those bounds. Regulators and recent commentaries have started to outline explainability and auditing expectations appropriate for such systems; our reliability analysis offers one concrete, measurable target.^25^

Our study complements emerging evaluations that move beyond static multiple choice to interactive, cost-sensitive diagnostic reasoning. In SDBench, a multiagent orchestrator (MAI-DxO) lifted model accuracy on NEJM Clinicopathological Case (CPC) cases to ∼80–86% while reducing diagnostic costs through role-structured debate, bias checking, and stewardship, techniques that specifically counter anchoring and premature closure. MedAgentBench similarly demonstrates that simple changes in output format and tool invocation can separate high performers from brittle ones, with the best success rates still leaving plenty headroom and with marked variation across task categories.^23^

We evaluated leading models on 200 rigorously standardized, temporally early cases derived from real errors and then stress-tested them with 2,200 controlled variants. This design isolates error correction ability under constraints and quantifies robustness to token changes. However, our cases, by design, feature eventual “teachable” outcomes and are not prevalence weighted; generalizability to routine emergency department and/or primary care presentations requires further study. We assessed single-turn judgment (with zeroshot prompts) rather than fully agentic, multiturn workflows; prior work suggests structured orchestration would likely improve correction rates and stability, particularly for models that benefited from comprehensive prompting in our sensitivity analysis.

Correcting one in two wrong initial diagnoses in high-uncertainty encounters would represent a meaningful step toward safer care. The work ahead is to turn this promise into practice by hardening systems against token-level instabilities, focusing on the disease categories that defeat both humans and machines at first pass, and deploying agentic, auditable workflows that amplify skepticism where it matters most, before error becomes outcome.

## Supporting information

Supplemental Attachment 1

## Data Availability

All data produced in the present study are available upon reasonable request to the authors and approval from our institution.

## Acknowledgement

We would like to thank and acknowledge Megan Clark for copy editing the final version of this article.

## Data Access

You can request data access in concordance with Johns Hopkins data sharing policy. Subset of the data is released as part of the benchmark in GitHub.

## References

1. Newman-Toker DE. Just how many diagnostic errors and harms are out there, really? It depends on how you count. BMJ Qual Saf [Internet] 2025 [cited 2026 Jan 9];34(6):355–60. Available from: https://qualitysafety.bmj.com/content/34/6/355

2. National Academy of Sciences. Improving Diagnosis in Healthcare [Internet]. 2015. Available from: http://www.nap.edu/catalog/21794 http://www.iom.edu/Reports.aspx

3. Newman-Toker DE, Nassery N, Schaffer AC, et al. Burden of serious harms from diagnostic error in the USA. BMJ Qual Saf [Internet] 2024 [cited 2024 Feb 4];33(2):109–20. Available from: https://qualitysafety.bmj.com/content/33/2/109

4. Newman-Toker DE, Wang Z, Zhu Y, et al. Rate of diagnostic errors and serious misdiagnosis-related harms for major vascular events, infections, and cancers: Toward a national incidence estimate using the “big Three.” Diagnosis [Internet] 2021 [cited 2023 Apr 4];8(1):67–84. Available from: https://www.degruyter.com/document/doi/10.1515/dx-2019-0104/html?lang=en

5. Hassoon A, Ng C, Lehmann H, et al. Computable phenotype for diagnostic error: developing the data schema for application of symptom-disease pair analysis of diagnostic error (SPADE). Diagnosis [Internet] 2024 [cited 2025 Jun 15];11(3):295–302. Available from: https://www.degruyterbrill.com/document/doi/10.1515/dx-2023-0138/html

6. McDuff D, Schaekermann M, Tu T, et al. Towards accurate differential diagnosis with large language models. Nature [Internet] 2025 [cited 2025 Sep 5];642(8067):451–7. Available from: https://www.nature.com/articles/s41586-025-08869-4

7. Liu X, Liu H, Yang G, et al. A generalist medical language model for disease diagnosis assistance. Nat Med [Internet] 2025 [cited 2025 Sep 5];31(3):932–42. Available from: https://www.nature.com/articles/s41591-024-03416-6

8. The Path to Medical Superintelligence | Microsoft AI [Internet]. [cited 2025 Sep 5];Available from: https://microsoft.ai/news/the-path-to-medical-superintelligence/

9. Zhou S, Xu Z, Zhang M, et al. Large Language Models for Disease Diagnosis: A Scoping Review. npj Artificial Intelligence [Internet] 2024 [cited 2025 Sep 5];1(1). Available from: https://arxiv.org/pdf/2409.00097

10. Newman-Toker DE, Sharfstein JM. The Role for Policy in AI-Assisted Medical Diagnosis. JAMA Health Forum [Internet] 2024 [cited 2026 Jan 9];5(4):e241339. Available from: https://pmc.ncbi.nlm.nih.gov/articles/PMC12419546/

11. Fanous A, Goldberg J, Agarwal A, et al. SycEval: Evaluating LLM Sycophancy. Proceedings of the AAAI/ACM Conference on AI, Ethics, and Society [Internet] 2025 [cited 2025 Nov 23];8(1):893– 900. Available from: https://ojs.aaai.org/index.php/AIES/article/view/36598

12. Liberman AL, Hassoon A, Fanai M, et al. Cerebrovascular disease hospitalizations following emergency department headache visits: A nested case–control study. Academic Emergency Medicine [Internet] 2022 [cited 2025 Jun 15];29(1):41–50. Available from:/doi/pdf/10.1111/acem.14353

13. DE N-T, SM P, S B, et al. Diagnostic Errors in the Emergency Department: A Systematic Review. Diagnostic Errors in the Emergency Department: A Systematic Review [Internet] 2022 [cited 2025 Jun 15];Available from: http://europepmc.org/books/NBK588118

14. Newman-Toker D, … NN-BQ&, 2023 undefined. Burden of serious harms from diagnostic error in the USA. qualitysafety.bmj.com DE Newman-Toker, N Nassery, AC Schaffer, CW Yu-Moe, GD Clemens, Z Wang, Y Zhu BMJ Quality & Safety, 2023•qualitysafety.bmj.com [Internet] [cited 2023 Oct 23];Available from: https://qualitysafety.bmj.com/content/early/2023/07/16/bmjqs-2021-014130?versioned=true

15. Singh H, Meyer AND, Thomas EJ. The frequency of diagnostic errors in outpatient care: estimations from three large observational studies involving US adult populations. BMJ Qual Saf [Internet] 2014 [cited 2023 May 31];23(9):727. Available from: /pmc/articles/PMC4145460/

16. Nassery N, Horberg MA, Rubenstein KB, et al. Antecedent treat-and-release diagnoses prior to sepsis hospitalization among adult emergency department patients: a look-back analysis employing insurance claims data using Symptom-Disease Pair Analysis of Diagnostic Error (SPADE) methodology. Diagnosis (Berl) [Internet] 2021 [cited 2021 Nov 6];8(4):469–78. Available from: http://www.ncbi.nlm.nih.gov/pubmed/33650389

17. Sharp AL, Pallegadda R, Baecker A, et al. Are Mental Health and Substance Use Disorders Risk Factors for Missed Acute Myocardial Infarction Diagnoses Among Chest Pain or Dyspnea Encounters in the Emergency Department? Ann Emerg Med [Internet] 2021 [cited 2021 Nov 6];Available from: http://www.ncbi.nlm.nih.gov/pubmed/34607739

18. Sharp AL, Baecker A, Nassery N, et al. Missed acute myocardial infarction in the emergency department-standardizing measurement of misdiagnosis-related harms using the SPADE method. Diagnosis 2021;8(2):177–86.

19. Mane KK, Rubenstein KB, Nassery N, et al. Diagnostic performance dashboards: Tracking diagnostic errors using big data. BMJ Qual Saf 2018;27(7):567–70.

20. Hassoon Ahmed, Austin Matt, Fanai Mehdi, et al. Big Data Dashboards to Measure Misdiagnosis-Related Harms – Visual Analytics to Identify Hospital Crossovers within and across Health Systems in Regional Health Information Exchange Datasets [Internet]. Baltimore: 2020. Available from: https://www.degruyter.com/view/journals/dx/5/4/article-peA59.xml

21. Newman-Toker DE, Schaffer AC, Yu-Moe CW, et al. Serious misdiagnosis-related harms in malpractice claims: The “Big Three” – vascular events, infections, and cancers. Diagnosis 2019;

22. Yao G, Zhang WJ, Zhu Y, et al. Comparing the accuracy of large language models and prompt engineering in diagnosing realworld cases. Int J Med Inform [Internet] 2025 [cited 2025 Nov 23];203. Available from: https://pubmed.ncbi.nlm.nih.gov/40617017/

23. Jiang Y, Black KC, Geng G, et al. MedAgentBench: A Virtual EHR Environment to Benchmark Medical LLM Agents. NEJM AI [Internet] 2025 [cited 2025 Sep 12];2(9). Available from: https://ai.nejm.org/doi/pdf/10.1056/AIdbp2500144

24. Wang W, Ma Z, Wang Z, et al. A Survey of LLM-based Agents in Medicine: How far are we from Baymax? 2025 [cited 2025 Sep 12];10345–59. Available from: https://aclanthology.org/2025.findings-acl.539/

25. Mesinovic M, Watkinson P, Zhu T. Explainability in the age of large language models for healthcare. Communications Engineering [Internet] 2025 [cited 2025 Sep 12];4(1):1–4. Available from: https://www.nature.com/articles/s44172-025-00453-y

26. Mehandru N, Miao BY, Almaraz ER, Sushil M, Butte AJ, Alaa A. Evaluating large language models as agents in the clinic. NPJ Digit Med [Internet] 2024 [cited 2025 Sep 12];7(1):1–3. Available from: https://www.nature.com/articles/s41746-024-01083-y

27. Qiu J, Lam K, Li G, et al. LLM-based agentic systems in medicine and healthcare. Nat Mach Intell [Internet] 2024 [cited 2025 Sep 12];6(12):1418–20. Available from: https://www.nature.com/articles/s42256-024-00944-1

28. The Path to Medical Superintelligence | Microsoft AI [Internet]. [cited 2025 Sep 12];Available from: https://microsoft.ai/news/the-path-to-medical-superintelligence/

29. Nori H, Daswani M, Kelly C, et al. Sequential Diagnosis with Language Models. 2025 [cited 2025 Sep 12];Available from: https://arxiv.org/pdf/2506.22405v1

