## Supplemental Attachment 1 for "Evaluating the AI Potential as a Safety Net for Diagnosis: A Novel Benchmark of Large Language Models in Correcting Diagnostic Errors"

Chief Complaint:
A 35-year-old female presents with chronic headaches, double vision, and occasional "pins and needles" sensation in her left arm for the past year.

History of Present Illness:
The patient reports frequent headaches, occurring approximately twice a week, which she attributes to stress from her work as an attorney. These headaches are often accompanied by brief episodes of double vision, which resolve spontaneously. Over the last six months, she has noticed intermittent tingling in her left arm, particularly when sitting at her desk for long periods. She denies weakness, balance issues, or other sensory changes.

Medical History:

Migraine headaches since her early 20s
Asthma, managed with as-needed inhalers
Medications:

Sumatriptan as needed for migraines
Albuterol inhaler
Family History:

Mother has migraines
No history of neurological or autoimmune diseases
Social History:

Lives with her partner; no children
Exercises regularly (yoga and swimming)
Non-smoker, drinks socially
Physical Examination:

Vital Signs: Blood pressure 122/80 mmHg, heart rate 74 bpm, respiratory rate 16 breaths per minute, temperature 36.9°C (98.4°F)
General: Well-appearing, no acute distress
Neurological Exam:
Cranial Nerves: Mildly reduced lateral gaze bilaterally during the exam; no nystagmus or ptosis
Motor Exam: 5/5 strength in all extremities
Sensory Exam: Decreased sensation to pinprick in the left forearm; other modalities intact
Reflexes: Symmetric and normal in upper and lower extremities
Cerebellar Exam: Normal coordination, Romberg negative

Clinical Course:
The patient is diagnosed with chronic migraine headaches with visual aura and possible repetitive strain injury (RSI) from desk work as the cause of her arm tingling. She is advised to use ergonomic tools and take regular breaks from typing.

Chief Complaint:
A 76-year-old woman presents to the clinic with a three-week history of involuntary, flinging movements of her right arm and leg. She describes the movements as abrupt, irregular, and interfering with daily activities, such as eating and dressing. She denies any pain, weakness, sensory changes, or other neurological symptoms.

History of Present Illness:
The patient reports that the movements began gradually and have progressively worsened. She initially attributed them to "nerve issues" but sought medical attention after dropping several items at home due to the severity of the movements. The movements persist throughout the day but are absent during sleep. She denies any recent infections, fevers, trauma, or changes in medication.

Medical History:

Hypertension, controlled with medication
Type 2 diabetes mellitus, poorly controlled (last HbA1c 8.2%)
Hyperlipidemia, treated with statins
No history of stroke, transient ischemic attack (TIA), or seizure disorders
Medications:

Metformin 500 mg twice daily
Amlodipine 10 mg daily
Atorvastatin 20 mg nightly
Family History:

Father had a stroke at age 80
Mother had hypertension, no history of movement disorders
Social History:

Retired teacher
Non-smoker, no alcohol or illicit drug use
Physical Examination:

Vital Signs: BP 138/82 mmHg, HR 80 bpm, RR 16 breaths/min, SpO2 97% on room air
General: Alert, cooperative, mildly distressed by movements
Neurological:
Cranial nerves II-XII intact
Motor: 5/5 strength in all extremities
Sensation: Normal to light touch and proprioception
Coordination: Normal finger-to-nose and heel-to-shin testing
Involuntary movements: Wild, irregular, nonrhythmic flinging movements of the right arm and leg, consistent with hemiballismus
Gait: Antalgic, with difficulty due to movements but no true ataxia
Speech: Normal, no dysarthria or aphasia
Laboratory Studies:

CBC: Normal
Random blood glucose: 16.2 mmol/L (normal: 3.9-11.1 mmol/L)
HbA1c: 8.5%
Electrolytes: Normal
Renal and liver function tests: Normal
Imaging:

Non-contrast head CT: No acute findings
MRI Brain (with DWI): Pending
Clinical Course:
The patient is diagnosed with hemiballismus, suspected secondary to poorly controlled diabetes mellitus causing dysfunction in the basal ganglia. She is started on glycemic control optimization and symptomatic management with clonazepam. Neurology follow-up is arranged for further evaluation.
