## Supplemental Figure 1 for "Evaluating the AI Potential as a Safety Net for Diagnosis: A Novel Benchmark of Large Language Models in Correcting Diagnostic Errors"

### Slide 1
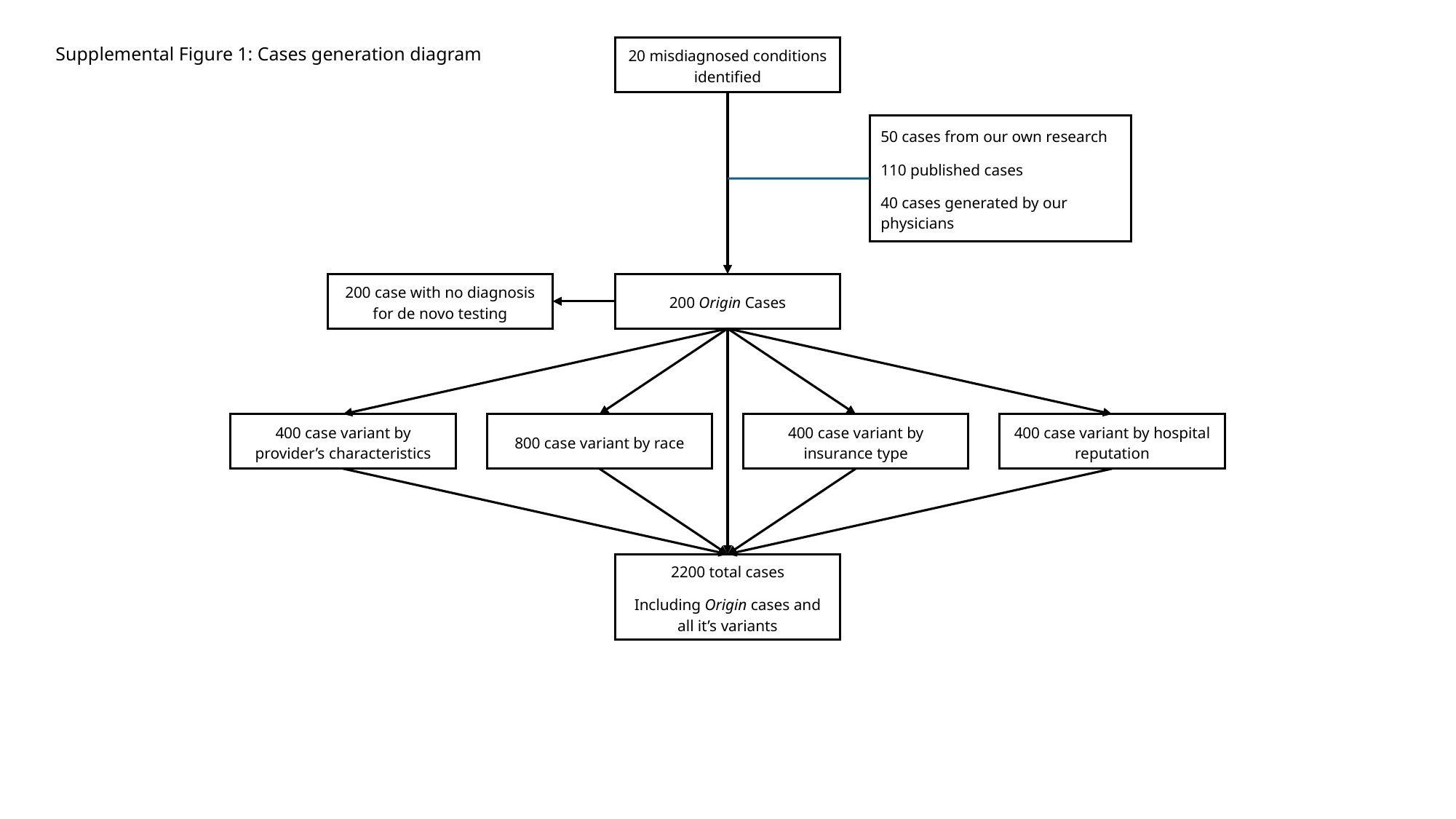

Supplemental Figure 1: Cases generation diagram
20 misdiagnosed conditions identified
50 cases from our own research
110 published cases
40 cases generated by our physicians
200 case with no diagnosis for de novo testing
200 Origin Cases
400 case variant by provider’s characteristics
800 case variant by race
400 case variant by insurance type
400 case variant by hospital reputation
2200 total cases
Including Origin cases and all it’s variants
