## Supplemental Table 1 for "Evaluating the AI Potential as a Safety Net for Diagnosis: A Novel Benchmark of Large Language Models in Correcting Diagnostic Errors"

Supplemental Table 1: LLMs Performance to Correct Diagnostic Errors by Diseases

| **Disease** | **female %** | **gemini_25_pro** | **claude_35** | **claude_4** | **opus_41** | **grok_4** | **claude_37** | **deepseek_r1** | **grok_3** | **gpt_5** | **gemini_2_flash** | **gpt_o1** | **gpt_45** | **grok_2** | **gpt_4o** | **nova_pro** | **deepseek_v3** |
| --- | --- | --- | --- | --- | --- | --- | --- | --- | --- | --- | --- | --- | --- | --- | --- | --- | --- |
| **Multiple sclerosis** | **60** | 90.0% | 90.0% | 70.0% | 80.0% | 80.0% | 90.0% | 80.0% | 90.0% | 80.0% | 70.0% | 80.0% | 80.0% | 80.0% | 70.0% | 60.0% | 40.0% |
| **Endocarditis** | **40** | 90.0% | 80.0% | 70.0% | 70.0% | 70.0% | 90.0% | 60.0% | 50.0% | 60.0% | 70.0% | 70.0% | 70.0% | 60.0% | 70.0% | 40.0% | 20.0% |
| **Melanoma** | **40** | 80.0% | 70.0% | 50.0% | 50.0% | 80.0% | 60.0% | 40.0% | 60.0% | 50.0% | 40.0% | 60.0% | 50.0% | 50.0% | 40.0% | 20.0% | 0.0% |
| **Male breast cancer** | **0** | 80.0% | 70.0% | 50.0% | 70.0% | 50.0% | 60.0% | 50.0% | 40.0% | 30.0% | 50.0% | 30.0% | 50.0% | 30.0% | 40.0% | 40.0% | 40.0% |
| **Multiple myeloma** | **30** | 80.0% | 60.0% | 70.0% | 70.0% | 60.0% | 70.0% | 50.0% | 70.0% | 50.0% | 50.0% | 70.0% | 60.0% | 70.0% | 20.0% | 70.0% | 40.0% |
| **Aortic dissection** | **30** | 70.0% | 70.0% | 70.0% | 60.0% | 60.0% | 60.0% | 60.0% | 50.0% | 60.0% | 60.0% | 50.0% | 40.0% | 40.0% | 40.0% | 40.0% | 40.0% |
| **Colorectal cancer** | **50** | 70.0% | 40.0% | 50.0% | 60.0% | 40.0% | 60.0% | 40.0% | 30.0% | 50.0% | 30.0% | 30.0% | 30.0% | 30.0% | 20.0% | 30.0% | 10.0% |
| **Pulmonary embolism** | **50** | 60.0% | 70.0% | 70.0% | 70.0% | 100.0% | 60.0% | 80.0% | 50.0% | 40.0% | 60.0% | 60.0% | 50.0% | 70.0% | 50.0% | 40.0% | 50.0% |
| **Deep venous thrombosis** | **30** | 60.0% | 50.0% | 70.0% | 50.0% | 60.0% | 60.0% | 60.0% | 50.0% | 60.0% | 60.0% | 50.0% | 60.0% | 40.0% | 30.0% | 30.0% | 30.0% |
| **Myocardial infarction** | **50** | 60.0% | 60.0% | 50.0% | 60.0% | 50.0% | 30.0% | 40.0% | 30.0% | 30.0% | 30.0% | 20.0% | 30.0% | 10.0% | 0.0% | 10.0% | 10.0% |
| **Stroke** | **40** | 50.0% | 60.0% | 50.0% | 50.0% | 60.0% | 50.0% | 50.0% | 50.0% | 80.0% | 50.0% | 50.0% | 50.0% | 40.0% | 30.0% | 20.0% | 30.0% |
| **Lung cancer** | **30** | 50.0% | 40.0% | 50.0% | 40.0% | 30.0% | 30.0% | 50.0% | 40.0% | 40.0% | 40.0% | 50.0% | 20.0% | 20.0% | 30.0% | 20.0% | 10.0% |
| **Sepsis** | **50** | 50.0% | 30.0% | 50.0% | 40.0% | 20.0% | 50.0% | 20.0% | 20.0% | 20.0% | 40.0% | 40.0% | 50.0% | 50.0% | 0.0% | 0.0% | 20.0% |
| **Prostate cancer** | **0** | 40.0% | 40.0% | 40.0% | 20.0% | 50.0% | 40.0% | 20.0% | 20.0% | 10.0% | 10.0% | 10.0% | 10.0% | 20.0% | 0.0% | 10.0% | 10.0% |
| **Female breast cancer** | **100** | 40.0% | 40.0% | 10.0% | 40.0% | 20.0% | 10.0% | 20.0% | 20.0% | 10.0% | 10.0% | 0.0% | 10.0% | 10.0% | 0.0% | 10.0% | 0.0% |
| **Pneumonia** | **50** | 30.0% | 60.0% | 40.0% | 50.0% | 50.0% | 30.0% | 30.0% | 40.0% | 40.0% | 30.0% | 40.0% | 30.0% | 30.0% | 30.0% | 30.0% | 20.0% |
| **Appendicitis** | **50** | 30.0% | 30.0% | 40.0% | 30.0% | 30.0% | 40.0% | 30.0% | 30.0% | 30.0% | 20.0% | 20.0% | 20.0% | 20.0% | 20.0% | 10.0% | 20.0% |
| **Spinal epidural abscess** | **40** | 30.0% | 0.0% | 10.0% | 10.0% | 20.0% | 10.0% | 10.0% | 0.0% | 10.0% | 10.0% | 10.0% | 0.0% | 10.0% | 0.0% | 0.0% | 10.0% |
| **Syphilis** | **40** | 20.0% | 10.0% | 30.0% | 0.0% | 10.0% | 0.0% | 20.0% | 0.0% | 10.0% | 0.0% | 0.0% | 10.0% | 10.0% | 0.0% | 0.0% | 0.0% |
| **Meningitis** | **40** | 20.0% | 0.0% | 0.0% | 20.0% | 0.0% | 10.0% | 10.0% | 20.0% | 0.0% | 20.0% | 0.0% | 10.0% | 0.0% | 0.0% | 0.0% | 0.0% |
| ***Average*** | *41* | *0.55* | *0.485* | *0.47* | *0.47* | *0.47* | *0.455* | *0.41* | *0.38* | *0.38* | *0.375* | *0.37* | *0.365* | *0.345* | *0.245* | *0.24* | *0.2* |
