## Supplemental Table 2 for "Evaluating the AI Potential as a Safety Net for Diagnosis: A Novel Benchmark of Large Language Models in Correcting Diagnostic Errors"

Supplemental Table 2: LLMs Performance by Token Insertion

| **Model (accuracy)** | ***Origine*** | **White** | **Black** | **Asian** | **Hispanic** | **Hopkins Hospital** | **Community Hospital** | **Hopkins-Trained** | **Community-Trained** | **Comprehensive Insurance** | **Basic Insurance** | **Runtime,**  **Mean (SD)*** |
| --- | --- | --- | --- | --- | --- | --- | --- | --- | --- | --- | --- | --- |
| Gemini 2.5 Pro | 0.55 | 0.50 | 0.56 | 0.56 | 0.54 | 0.53 | 0.55 | 0.53 | 0.56 | 0.50 | 0.53 | 134.3(18.5) |
| Claude sonnet 4 | 0.47 | 0.51 | 0.50 | 0.52 | 0.50 | 0.48 | 0.51 | 0.49 | 0.49 | 0.48 | 0.49 | 4.8(0.3) |
| Claude Opus 4.1* | 0.47 | 0.46 | 0.51 | 0.48 | 0.52 | 0.49 | 0.48 | 0.49 | 0.52 | 0.47 | 0.50 | 38.7(2.5) |
| Claude 3.5 | 0.49 | 0.48 | 0.51 | 0.49 | 0.48 | 0.48 | 0.49 | 0.45 | 0.49 | 0.48 | 0.49 | 6.3(1.3) |
| Grok 4 | 0.47 | 0.47 | 0.45 | 0.45 | 0.49 | 0.47 | 0.46 | 0.44 | 0.45 | 0.47 | 0.46 | 114.3(20.5) |
| Deepseek R1 | 0.41 | 0.42 | 0.46 | 0.46 | 0.43 | 0.45 | 0.41 | 0.46 | 0.43 | 0.44 | 0.42 | 66.9(10.4) |
| GPT 5 | 0.38 | 0.37 | 0.39 | 0.38 | 0.36 | 0.39 | 0.38 | 0.38 | 0.41 | 0.39 | 0.38 | 248.0(4.7) |
| Grok3 | 0.38 | 0.39 | 0.41 | 0.39 | 0.39 | 0.38 | 0.38 | 0.38 | 0.39 | 0.36 | 0.36 | 2.1(0.9) |
| O1 | 0.37 | 0.36 | 0.36 | 0.36 | 0.38 | 0.34 | 0.35 | 0.34 | 0.35 | 0.31 | 0.32 | 61.7(19.9) |
| Amazon Nova Pro | 0.24 | 0.27 | 0.28 | 0.28 | 0.28 | 0.26 | 0.28 | 0.26 | 0.28 | 0.25 | 0.26 | 4.2(0.8) |
| Average | 0.42 | 0.42 | 0.44 | 0.43 | 0.44 | 0.42 | 0.43 | 0.42 | 0.43 | 0.41 | 0.42 | 68.1 |

* Extended thinking (max_reasoning tokens = 1024). Runtime is average per 200 case batches measured in minutes
