## Supplemental Table 3 for "Evaluating the AI Potential as a Safety Net for Diagnosis: A Novel Benchmark of Large Language Models in Correcting Diagnostic Errors"

Supplemental Table 1: LLMs Reliability Measures by Tokens Insertion

| Model | Mean Token Performance | Standard Deviation | Coefficient of Variation | Signal to Noise Ratio | Normalized Range | Raw Score | Composite Reliability |
| --- | --- | --- | --- | --- | --- | --- | --- |
| Gemini 2.5 Pro- formal | 0.53 | 0.02 | 0.04 | 24.12 | 0.11 | 0.15 | 0.38 |
| Claude sonnet 4 | 0.49 | 0.01 | 0.03 | 36.83 | 0.09 | 0.12 | 0.52 |
| Claude Opus 4.1 * | 0.49 | 0.02 | 0.04 | 23.88 | 0.12 | 0.16 | 0.34 |
| Claude 3.5 | 0.48 | 0.01 | 0.03 | 34.24 | 0.11 | 0.14 | 0.42 |
| Grok 4 | 0.46 | 0.02 | 0.03 | 30.28 | 0.11 | 0.14 | 0.43 |
| Deepseek R1 | 0.44 | 0.02 | 0.04 | 22.98 | 0.13 | 0.17 | 0.32 |
| GPT 5 - medium | 0.38 | 0.01 | 0.03 | 31.12 | 0.12 | 0.15 | 0.40 |
| Grok3 | 0.38 | 0.01 | 0.04 | 28.34 | 0.12 | 0.15 | 0.38 |
| O1 | 0.34 | 0.02 | 0.06 | 16.82 | 0.19 | 0.25 | 0.00 |
| Amazon Nova Pro | 0.27 | 0.01 | 0.04 | 22.66 | 0.13 | 0.18 | 0.29 |

* Extended thinking (max_reasoning tokens = 1024)
